# Motor signatures in digitized cognitive and memory tests enhances characterization of Parkinson’s disease

**DOI:** 10.1101/2022.03.28.22272824

**Authors:** Jihye Ryu, Elizabeth B Torres

## Abstract

**Background:** Although there is a growing interest in using wearable sensors to characterize movement disorders, there is a lack of methodology for developing clinically interpretable kinematics biomarkers. Such digital biomarkers would provide a more objective diagnosis, capturing finer degrees of motor deficits, while retaining the information of traditional clinical tests.

**Objectives:** We aim at digitizing traditional tests of cognitive and memory performance to derive motor biometrics of pen-strokes and voice, thereby complementing clinical tests with objective criteria, while enhancing the overall motor characterization of Parkinson’s disease (PD).

**Methods:** 35 participants including patients with PD, healthy young and age-matched controls performed a series of drawing and memory tasks, while their pen movement and voice were digitized. We examined the moment-to-moment variability of time-series reflecting the pen speed and voice amplitude.

**Results:** The stochastic signatures of the fluctuations in pen drawing speed and voice amplitude of patients with PD show lower noise-to-signal ratio compared to those derived from the younger and age-matched neurotypical controls. It appears that contact motions of the pen strokes on the tablet evokes sensory feedback for more immediate and predictable control in PD, compared to controls, while voice amplitude loses its neurotypical richness.

**Conclusions:** We offer new standardized data types and analytics to help advance our understanding of hidden motor aspects of cognitive and memory clinical assays commonly used in Parkinson’s disease.

## 1. Introduction

The advent of wearable biosensors and their use in research throughout the last decade, spurred the use of kinematic parameters to characterize symptoms of Parkinson’s disease (PD) [1-6]. For example, a search on app.dimensions.ai using the words kinematic biomarkers and Parkinson’s disease yielded 2,990 peer-reviewed papers since 2016, that we can visualize using some features of the VOS Viewer software for bibliographic analyses [7]. Figures 1A-B show clusters of main labs using kinematics for analyses of PD symptoms since 2016 to present, and Figures 1C-D show the main peer-reviewed journals publishing this type of research since 2013.

**Figure 1.**
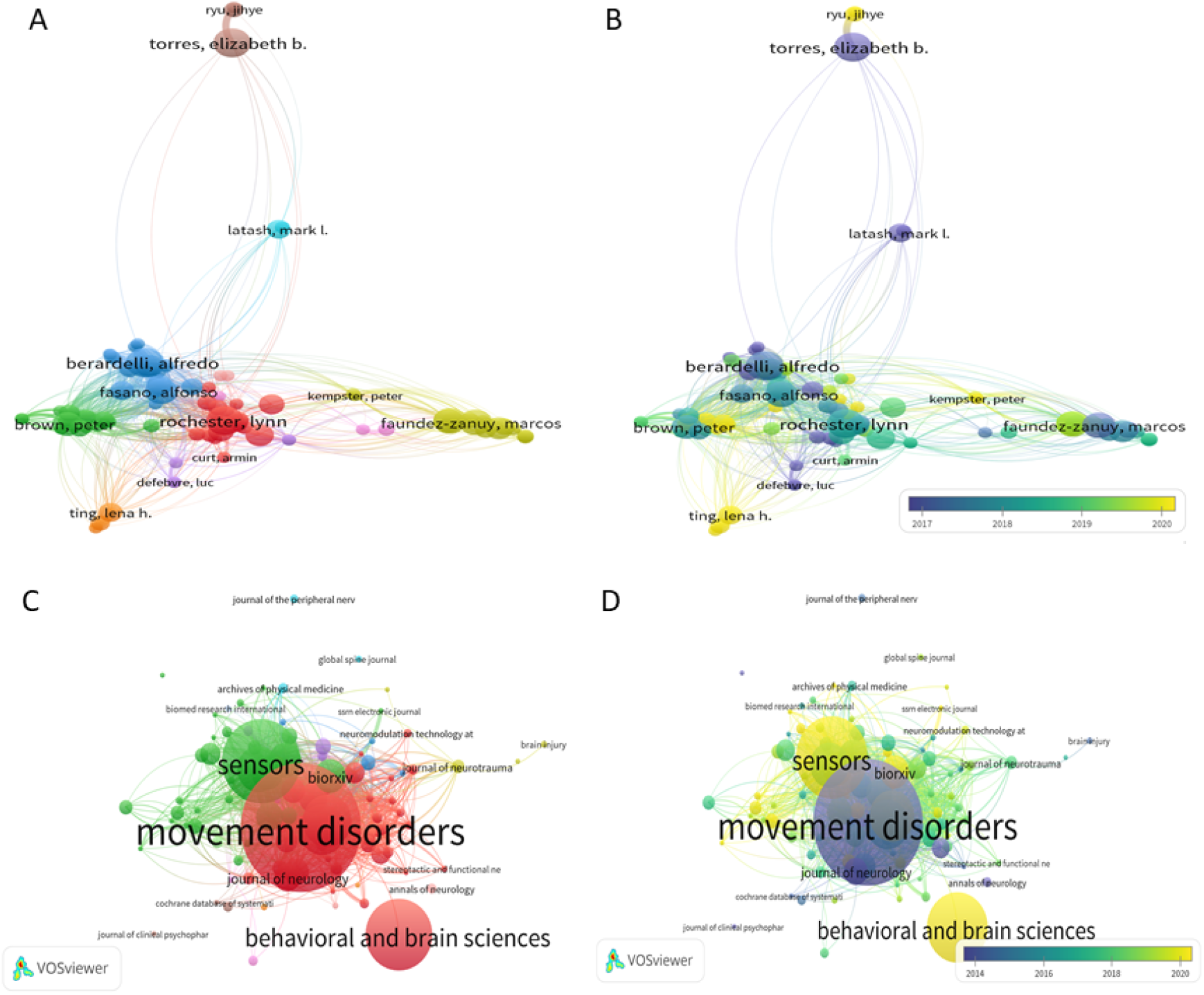
Bibliographic analyses of publications using kinematic parameters to characterize symptoms of PD reveal clusters by research labs over time (A-B) and publication journals (D-E) over time.

In the past, criteria to assess PD was more reliant on observation and pencil and paper inventories that clinicians have perfected over time, under best-practice standards that include training, certification, and scientific exchange to create reliable tools. Notwithstanding their rigor and ease of use for scalability, relying exclusively on observation misses an opportunity to see beyond the limits of the naked eye. With new off-the-shelf biosensors that have high sampling resolution and research-grade quality, we are now well positioned to complement the experienced clinical eye with digital data, to produce interpretable digital biomarkers. This new concept emerges from digitizing existing clinical tests, e.g., the Universal Parkinson’s Disease Rating Scale (UPDRS) [8] and other cognitive tests like the Montreal Cognitive Assessment (MOCA) [8] commonly used at clinical settings. Such digital characterization of various aspects of the disorder are easy to do because wearable biosensors are non-obtrusive and can be easily placed on the person’s body, clothing or even collect data streams of motion and voice as the person performs the tasks that are typically administered at clinical settings, or over the internet, e.g., via zoom or using other telemedicine means.

Biosensor’s data offers many advantages, as it increases the level of granularity of the motor phenomena and enables us to learn more about the internal physiological activities of the person’s nervous systems. However, analytical techniques often rely on summary statistics under theoretical assumptions of linearity, normality, and stationarity in the data, when, empirically, the streams of analogue data that we collect can change in highly non-linear ways, not be normally distributed and be non-stationary, within the time scale of hours or even minutes. Indeed, we have found that kinematic parameters change nonlinearly [4, 6], tend to not distribute normally [9, 10] and shift signatures of variability from moment to moment in ways that allow differentiation between patients with PD and controls [5].

For all these reasons, we created a new data type (coined micro-movement spikes, MMS) that permits us to provide an empirically informed characterization of the movement phenomena across different parameter spaces and data stream modalities. This is done by combining information from different data streams (e.g., voice and motion) during typical clinical assessments (Benson complex figure, trail making test, clock drawing) and memory tests. These digitized clinical assays can be very revealing of the personalized signatures and of the cohort’s signatures and trends. Here we employ these new methods and analyze motor signatures from voice and motions during commonly performed cognitive and memory tasks that are part of the clinical assessments used in PD.

## 2. Methods

### 2.1. Participants

A total of 35 participants partook in this study. Among these, 18 participants (age 28-77) were recruited from the Robert Woodrow Johnson Medical Center at Rutgers University or Clinical Trials website (clinicaltrials.gov). We had 11 patients with PD and 5 age-matched neurotypical participants (see Table A1), and 17 undergraduate students (age 18-35) recruited as young healthy controls from the Rutgers University human subject pool system. All participants provided informed consent, which was approved by the Rutgers University Institutional Review Board.

From a series of experiments [11], we report a subset, namely, the drawing and memory tasks, from which we analyzed the pen motion and voice data respectively. All but two participants performed these in-person, while two of the age-matched controls performed only the memory tasks remotely via Zoom (San Jose, California).

### 2.2. Experimental procedure

The participant completed two tasks – drawing and memory (Figure 2).

**Figure 2.**
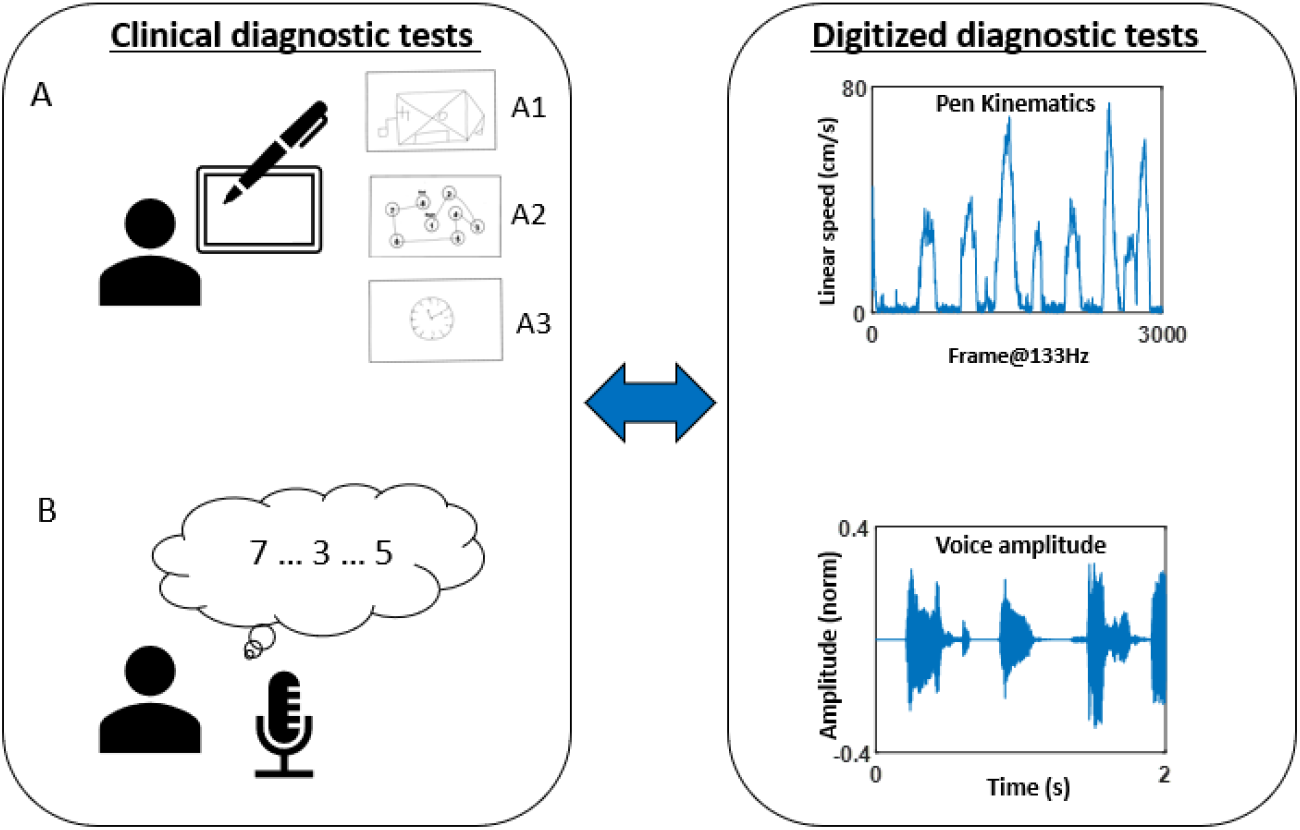
(Left) Clinical diagnostic tests of drawing and memory tasks were performed. (Right) These were digitized and assessed through kinematic analyses of the pen motion during the drawing task, and audio analyses of the voice during the memory task.

For the drawing task, the participant used a digitizing pen and tablet (Wacom, Japan) to complete seven drawing tasks, which are subtasks of standardized clinical diagnostic tests. Specifically, the tasks were to 1) copy a Benson Complex Figure [12] (denoted as “BCopy” in Figure 2A.A1), 2) connect 8 circles in numerical order (denoted as “TrailA” in Figure 2A.A2), 3) connect 25 circles in numerical order, 4) connect 8 circles composed of 4 numbers and 4 letters in alpha-numerical order (denoted as “TrailB(s)”), 5) connect circles of 13 numbers and 12 letters in alpha-numerical order (denoted as “TrailB”), 6) draw an analog clock and set the time to 10 past 11(denoted as “Clock” in Figure 2A.A3), and 7) draw the Benson Complex Figure from memory (denoted as “BDelay). Motions from the digital pen was recorded with the software MovAlyzer (Neuroscript; Tempe, AZ), sampling the position of the pen tip at 133Hz.

For the memory task, the researcher recited a string of numbers (ranging from 2 to 9 digits), and the participant repeated them in the same order for the forward memory task, and in a reverse order for the backward memory task [13]. The researcher continued until the participant failed to repeat them in two consecutive trials. The audio of the participant’s voice was recorded with a microphone sampled at 48,000 Hz. For the two participants who completed the task remotely, the voice was recorded from an audio clip provided by Zoom (San Jose, California), sampled at 44,100Hz.

### 2.3. Preprocessing Methods

The audio data collected during the memory tasks were continuously recorded while the researcher and participant alternated saying a string of numbers. We manually removed the researcher’s voice from the audio using Audacity (open source software; version 2.3.1), and glued the participant’s voice from different trials. Then, the audio was decomposed using the Gammatone filter [14, 15] in MIR toolbox [16] (Appendix Figure A1.A-C). The Gammatone function is defined in the time domain by its impulse response, and is known to simulate the response of the basilar membrane [14]. The decomposed audio was then enveloped in the MIR toolbox (Appendix Figure A1.D). This produced 10 different enveloped audio amplitudes, which we examined separately to find which range of band was most informative to characterize PD. While we found a similar pattern across all bands, we found the 8^th^ band filter (ranging 4000-7000Hz, centered at 5600Hz) to be the most informative. For that reason, we present the results based on this filtered enveloped audio data.

Note, 2 participant’s data were sampled at a different rate (44,100Hz) from the rest of the participants (48,000Hz). We analyzed the data by down sampling them to the be at the same rate but did not find much difference in the overall pattern that we would describe in the later section. For that reason, we preserved the audio data at their actual sampling rates in the analysis for everyone.

### 2.4. Data Analysis

#### 2.4.1. Stochasticity of pen movement during drawing task

The positional trajectory of the pen tip was registered (Figure 3A) and its linear speed was computed (Figure 3B). Note, linear speed was computed for the entire duration, which includes the times when the pen was lifted from the tablet (see Figure A2 for the percentage of pen lifting). The peaks and minima were extracted from the linear speed time series (denoted in red and cyan, respectively in Figure 3B), which were then converted into a unitless micromovement spike (MMS) data (Figure 3C) [17, 18]. These standardized spike amplitudes were computed using the Equation 1 below:

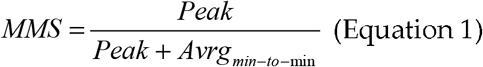

**Figure 3.**
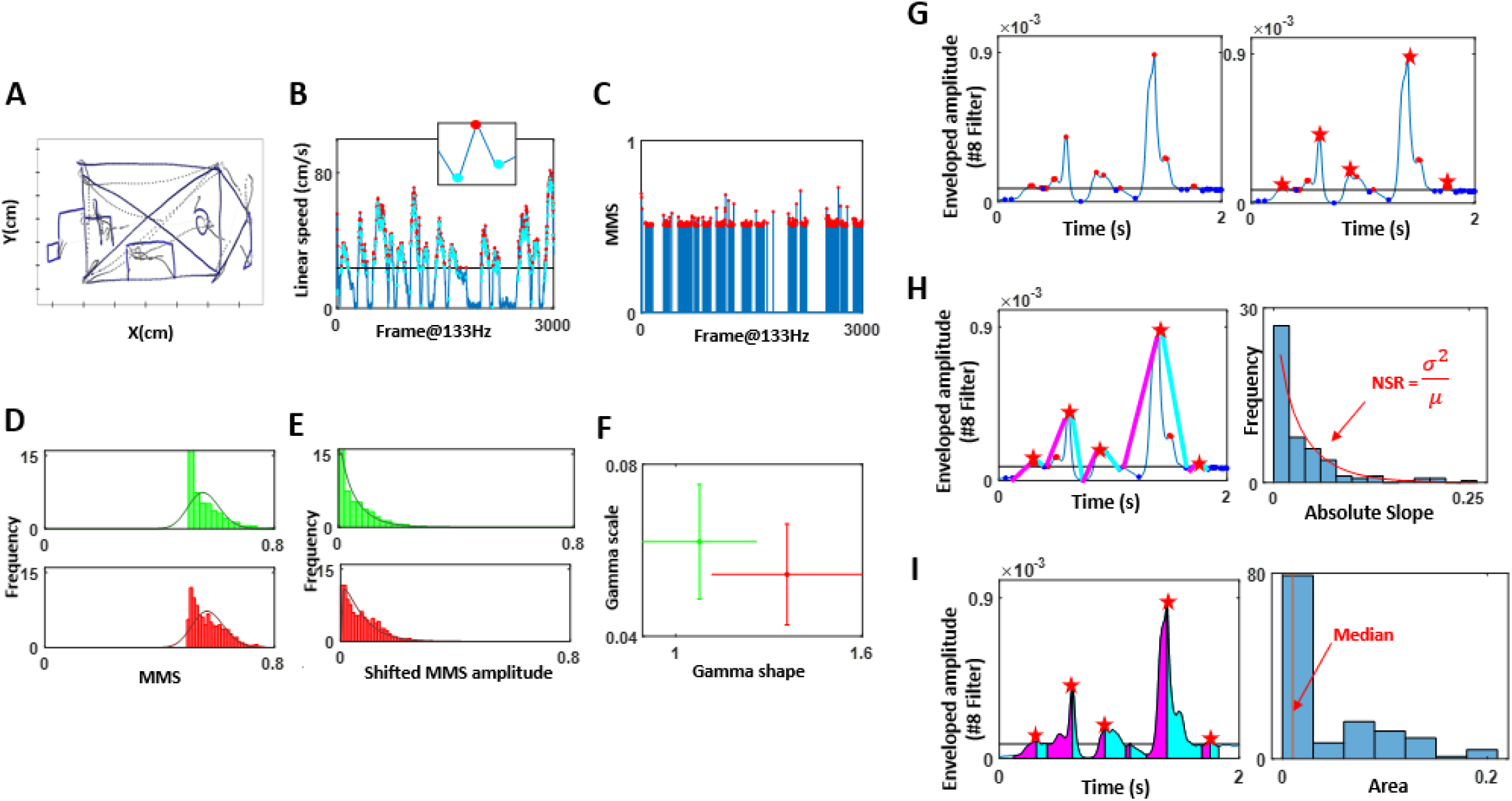
(A-F) Sample methods of data acquisition and analyses for drawing motions. (G-I) Audio analyses using attack and decay segments of the enveloped signal.

Note, we only extracted the MMS values with raw peak amplitudes exceeding the median linear speed (denoted in black horizontal line in Figure 3B), as these high peak amplitudes were most informative to characterize PD motions. The inset in Figure 3B zooms into a speed peak (red) and its surrounding local minima (cyan), which are the elements of computing MMS (Equation 1). This equation scales out the anatomical differences that would otherwise introduce allometric effects on the analyses [19].

To observe the stochasticity of the pen movement, the MMS (standardized spikes peaks) were plotted on a histogram (Figure 3D), and showed to be concentrated at 0.5, exhibiting a shifted exponential-like distribution shape. To allow a good fit to a family of distributions, the MMS amplitude values were shifted by subtracting 0.5 uniformly (Figure 3E). As a result, the histogram of such shifted MMS was best fit to a Gamma PDF using maximum likelihood estimation (MLE). To characterize individual variability in PD, the fitted Gamma (shape, scale) parameters were plotted for each person, with 95% confidence intervals on a Gamma parameter plane, e.g., two participants in Figure 3F in red and green. The stochastic signatures of these pen motions were compared against the clinical scores obtained from the drawings. Specifically, for the task of copying the Benson complex figure (BCopy) and drawing it from memory (BDelay), the scores were based on the accuracy of the drawing and ranged from 1 to 17. The trail making tests (TrailA, TrailB) were scored based on the time (seconds) that it took to complete the task. Because the sample test of Trail A was short (less than 5 seconds for most participants), we did not include this in our analysis. Lastly, the clock drawing task (Clock) was scored based on the accuracy of the drawing and ranged from 0 to 3.

It is noteworthy that fitting the Gamma PDF allows us to examine a wide range of probability distributions, from exponential to Gaussian [10, 17, 20], and to interpret the stochastic results with mathematical meaning. Specifically, the Gamma shape parameter ranges from value of 1 to above 100. The special case of shape = 1 corresponds to the memoryless exponential distribution, whereby immediate future events are equally probable, and no predictive pattern drives the system. This is the case when information is random and exists in “the here and now”. As the shape value increases, the distribution shifts from skewed (with heavy tails) to symmetric, highly predictive (Gaussian) patterns. Empirically, we have previously seen high randomness in the motor stochastic signatures of patients with PD [5], and interpreted that the type of kinesthetic (reafferent) feedback that such signatures reveal is less than ideal to predict the consequences of (efferent) action signals, and to compensate for sensory-motor transduction and transmission delays in the system [6, 17, 21].

The other parameter, the Gamma scale, is equivalent to the noise-to-signal ratio (NSR) (i.e., variance over mean, because given a as the Gamma shape and b as the Gamma scale, the Gamma mean is a*b and the Gamma variance is a*b^2^ such that their noise to signal ratio is b, the Gamma scale). Empirical work relating the log-log parameters show an inverse relationship between the log NSR (log Gamma scale) and the log Gamma shape. Gaussian regimes correspond to low NSR, and we empirically find these in healthy controls, yielding highly interpretable value to our approach. Indeed prior studies have demonstrated NSR to be high among patient populations (e.g., ASD [22], Parkinson’s disease [23], schizophrenia [24]) using their kinematics during naturalistic motions, and be susceptible to change with higher motor intent (e.g., pointing [25-27]).

#### 2.4.2. Stochasticity of attack and decay phases of speech

The enveloped audio filtered at 4,000-7,000Hz (centered at 5,600Hz) was examined by segmenting the data by attack (upwards) and decay (downwards) phases. First, minima and maxima were extracted, with a criterion that maxima be above the median amplitude’s values, and minima be below the median (Figure 3G, left). This is because we wanted to focus on meaningful maxima and minima that were most likely produced by actual speech, and ignore pauses and non-speech segments (e.g., breathing, sighing). Since there were cases where multiple maxima were present between 2 minima, and because our analytics involved segmenting the attack and decay phases (where attack phase would start from the local minimum to the subsequent local maximum, and decay phase start from the local maximum to the subsequent local minimum), it was necessary to have only 1 maximum between 2 minima. For that reason, when multiple local maxima were present between 2 adjacent minima, we chose the maximum (denoted by red star in Figure 3G, right) as the start and end points of attack and decay phases.

Then, we computed the attack and decay slopes. The attack slope was computed by taking the slope between the local minimum and the subsequent maximum (denoted by magenta line in Figure 3H), and the decay slope between the maximum and subsequent minimum (denoted by the cyan line). The absolute slopes were then plotted on a histogram and their NSR (i.e., fitted Gamma PDF scale parameter) were compared between PD and other groups. We also computed the area under the curve during the attack and decay phases, by taking the Riemann sum during the two phases as shown in magenta and cyan respectively in Figure 3I. Then the stochasticity of these areas was plotted on a histogram and the median values were examined to characterize PD against other groups. Finally, the stochasticity of these attack and decay phases were examined against the clinical scores of the memory task, which was the average of the longest digits correctly recited during the forward and backward memory tasks.

Note, we tested with other measures (e.g., NSR of area under the curve, median of absolute slopes), but found the NSR of absolute slopes, and median of area under the curve to best characterize and differentiate PD from controls. We also tested these parameters separately for the forward and backward memory tasks but did not find much difference between the two. For that reason, we combined the audio from the forward and backward tasks for the analysis.

## 3. Results

### 3.1. Patients with PD show lower noise-to-signal ratio (NSR) in their pen motion

The fitted Gamma parameters alongside the actual drawing scores were plotted for each participant, for each drawing task in Figure 4A. This figure spans a parameter space amenable to detect self-emerging patterns across the cohort. We also set landmarks of the most severe case of PD according to the clinical scores (upright triangle) vs. the least severe case (inverted triangle).

**Figure 4.**
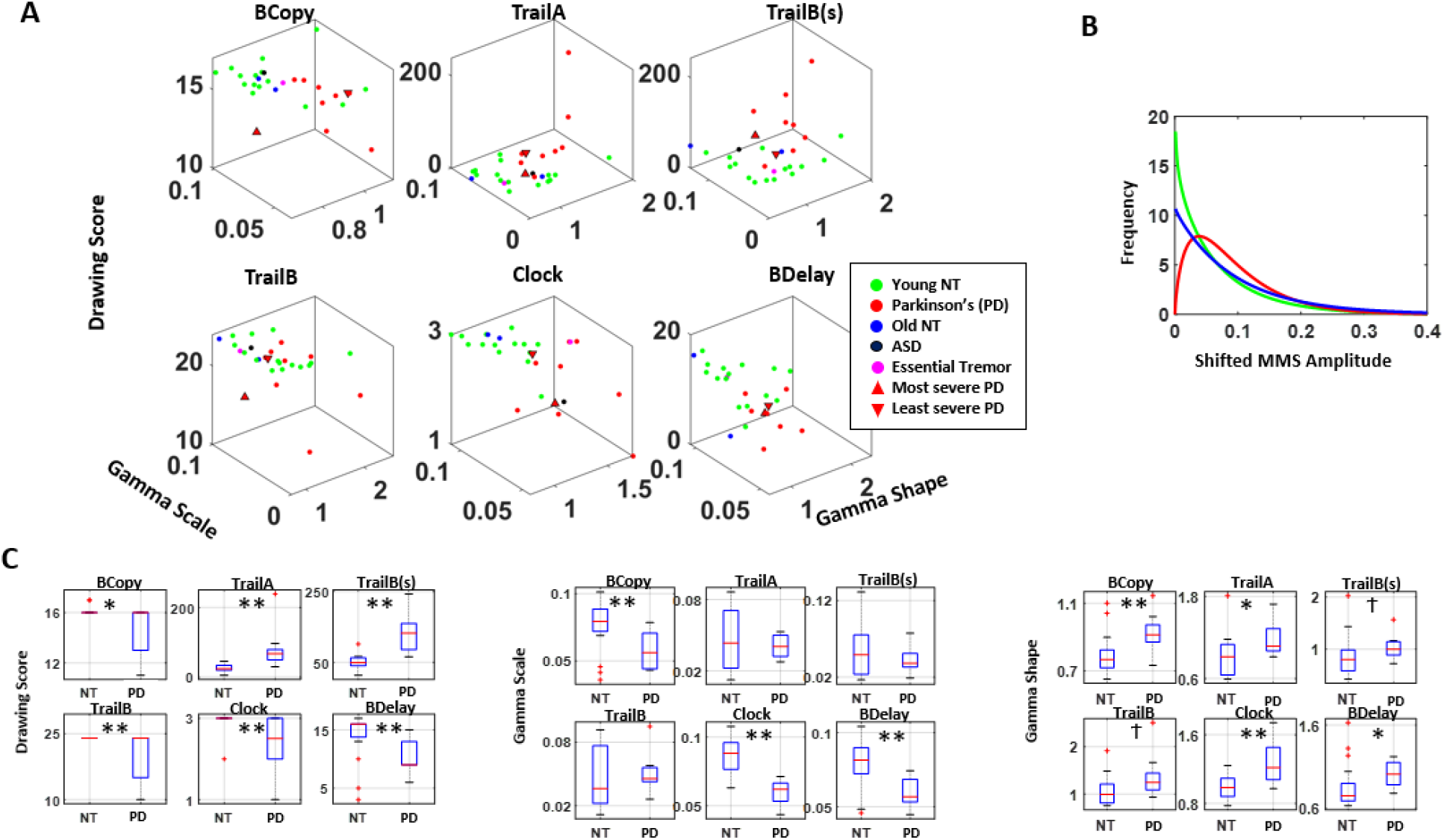
Combining Gamma parameters scale, shape and drawing scores separates the stochastic signatures derived from the drawing motions of young controls (denoted as NT) and PD. ** p<0.01, * p<0.05, t p<0.1

Across all drawings, patients with PD showed lower drawing scores than the young healthy cohort (NT), (Figure 4C left, Table A2). When we examine the stochasticity of pen motion, PD tend to have a lower scale and higher shape parameter than NT (Figure 4 middle, right, Table A2). The single participant with ASD and the patient with Essential Tremor did not show noticeable difference in their stochasticity, compared to NT. The separation between PD an NT is most pronounced during the clock and the Benson complex figure task (both BCopy and BDelay), which involve drawing on a blank sheet of paper, which allows more freedom in motion. These tasks contrast with others, where participants are given a paper with some form of figures already printed on it (e.g., circles with letters and/or numbers), and thus have less freedom in their pen motion. We also analyzed this separately between older NT and the PD, and still found similar patterns with reduced difference in the stochasticity as shown in Figure A3, thus confirming that the observed pattern between PD and NT is not solely due to age.

When we examined the PDF, the typical patient with PD tends to have different PDF than NT, as shown in Figure 4B. This alludes to the nature of contact control (control while body is in contact with an object) among individuals with PD, where they tend to exhibit lower NSR in their hand motion variability, especially in cases where movement traces are not prescribed (as in the clock and Benson figure tasks). This can be a useful addition to the current drawing diagnostic tests, because certain drawing test scores (particularly the clock and Benson figure copy) tend to be too coarse to differentiate participants, as most end up reaching the ceiling score.

### 3.2. Patients with PD show lower change and NSR in voice amplitude

Based on the enveloped audio filtered at 4,000-7,000Hz, the NSR of the distribution of absolute attack and decay slopes, and the median area under the curve during the attack and decay phases, alongside the actual memory scores obtained during the memory task are plotted for each participant in Figure 5A.

**Figure 5.**
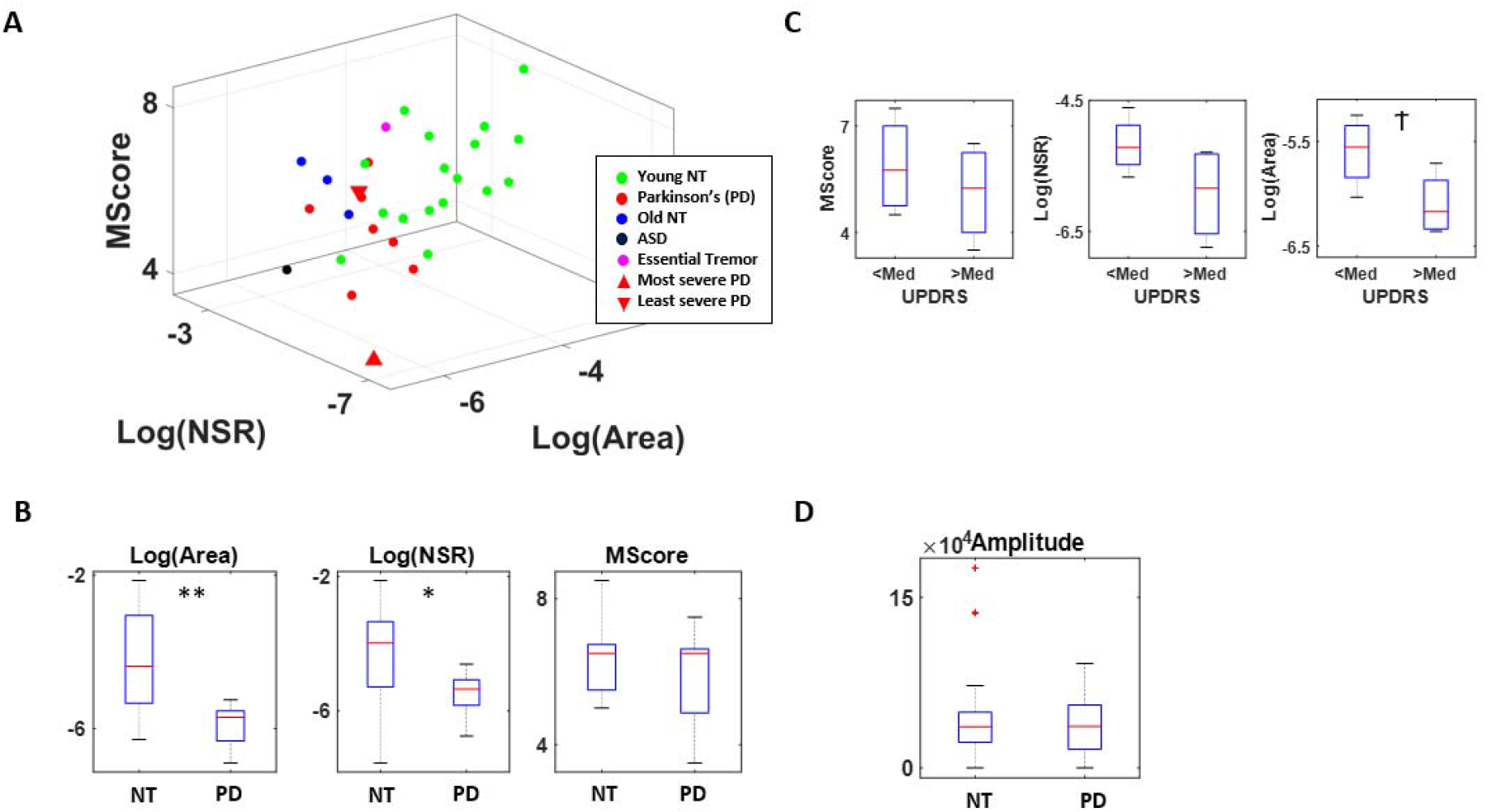
Combining attack and decay parameters (NSR, Area) and memory scores (MScore) separates the stochastic signatures derived from the voice amplitudes of young controls (denoted as NT) and those with PD. **p<0.01, * p<0.05, t p<0.1

The memory score itself does not show statistical difference between PD and NT, although patients with PD tend to show lower scores (χ(1,27) = 0.31, p=.58). On the other hand, there is a clear separation between the two groups when comparing the area under the curve (χ(1,27) = 9.98, p<0.01) and the NSR of absolute slope distribution (χ(1,27) = 4.70, p=0.03) (Figure 5B). Specifically, patients with PD tend to show a smaller area under the curve and lower NSR in their distribution, and this pattern is maintained when comparing patients with PD against their older NT counterparts (for area under curve, χ(1,10) = 5.34, p=0.02; for NSR of slope distribution χ(1,10) = 6.23, p=0.01) confirming that this difference is not due to age (Figure A4).

When we examine this question only among the PD group and compare those with higher severity (with UPDRS above the median) against those with less severity (with UPDRS below the median) as shown in Figure 5C, we find a similar pattern, suggesting that this measure not only characterizes PD but also reflects the severity of the disorder. To exclude such separation to be due to difference in voice volume, we also compared the voice amplitude between PD and NT and found the two groups to have a similar range of volume in their voices (Figure 5D). We also tested such comparison across different audio bands, and found similar patterns, but most pronounced at frequency 4,000-7,000Hz (Figure A5), which is what we show in Figure 5.

Overall, these findings imply that the variability in attack and decay phases of the voice from patients with PD are stochastically different, such that changes in voice amplitude (particularly within 4,000-7,000Hz range) is smaller and less variable than the NT. These characteristics may be useful additions to cognitive tests, as accompanying deficits in the neuromotor aspects of PD.

## 4. Discussion

This work extends the fast-growing body of knowledge involving kinematic analyses of motion patterns from patients with PD and controls across different age groups. We combined data from drawing motion trajectories of a pen on a tablet and the participant’s voice, to provide a set of methods amenable to use in the lab, clinic or remotely over zoom, as the person undergoes traditional clinical evaluations or telemedicine visits. These methods digitized typically administered tests such as parts of the MOCA, assessing memory and cognitive status of the person, and took advantage of the (hidden) motor components of these tests. Using a new data type, the MMS and a combination of non-parametric statistical analyses and data-driven stochastic analyses, we were able to demonstrate clear separations between patients with PD and controls.

We found that in the drawing tasks, patients with PD manifest lower values of the Gamma scale and higher values of the Gamma shape derived from the MMS of their drawing speed. Their motions are far more controlled and predictable than those of neurotypical young and older participants. The latter are well characterized by fluctuations in speed that distribute exponentially, in contrast to those of PD who had speed fluctuations distributed in skewed to symmetric shape ranges.

This result contrasts with the signatures of pointing motions to visual targets, whereby higher NSR and exponentially distributed fluctuations prevail in PD [6]. We attribute these differences to the pen’s contact with the tablet, providing continuous feedback to the motor performance. In PD, motor control is already thought to be under deliberate supervision of the motor systems [28], partly accounting for the bradykinetic motions known to characterize their motor performance [4]. Unlike in pointing motions, where no contact surface provides feedback and the feedback is rather kinesthetic from the fluctuations of the motion trajectories themselves [6], here we can appreciate this overt supervision with significantly lower speeds and highly controlled fluctuations of the drawing traces, in relation to controls. This result is important because in addition to offering new ways to use e.g., the MOCA tests to assess motor components, it suggests a new possible target for treatment. Namely, a possible new way to directly provide feedback to the motor systems is through contact motions, that through this pressure-based reafferent channel could aim at bringing the NSR of their fluctuations in traces’ speed, to levels comparable to those of controls.

Using the voice data, we also found ways to separate patients with PD from controls and demonstrated the differences in parameter spaces for characterizing PD. These methods of data acquisition and analyses are amenable to use with zoom, for remote research and adds to our arsenal of instruments that the recent pandemic generated to enable continuity of our research programs using off-the-shelf means.

The methods presented in this work are personalized, as they reflect the person’s stochastic signatures of motor variability, uniquely localizing everyone in the cohort on a parameter space. Furthermore, interpretation of the data from the group is possible as clusters self-emerge within the parameter space, often automatically separating the patients from the neurotypical controls. As the data are standardized and possible allometric effects scaled out, the MMS derived from the moment-to-moment fluctuations in signal amplitude (e.g., speed or other derived parameters like area under the attack-decay curve), we can add other patient populations with motor issues. We can aim at automatically stratifying the heterogeneous spectrum of clinically-defined disorders and identifying which parameters may optimally separate diverse disorders of the nervous systems. Using motoric components hidden in these minute fluctuations – that we tend to throw away as superfluous, gross data – we made good use of these standardized fluctuations, offering new ways to leverage existing cognitive and memory inventories broadly used by the field of clinical and basic research.

In summary, we offer new standardized data types and analytics to help advance the clinical and applied research of Parkinson’s disease.

## Data Availability

All data produced in the present study are available upon reasonable request to the authors

## Acknowledgment

Research assistants: Christina Wilson, Nishad Nalgundwar, Shaniqua Lewis

## Authors’ Roles

1. Research project: JR, EBT conception; JR, EBT organization; JR, EBT execution.
2. Statistical Analysis: JR, EBT design; JR, EBT execution; JR, EBT review and critique.
3. Manuscript: JR, EBT writing of the first draft; JR, EBT review and critique.

## Appendix

**Figure A1.**
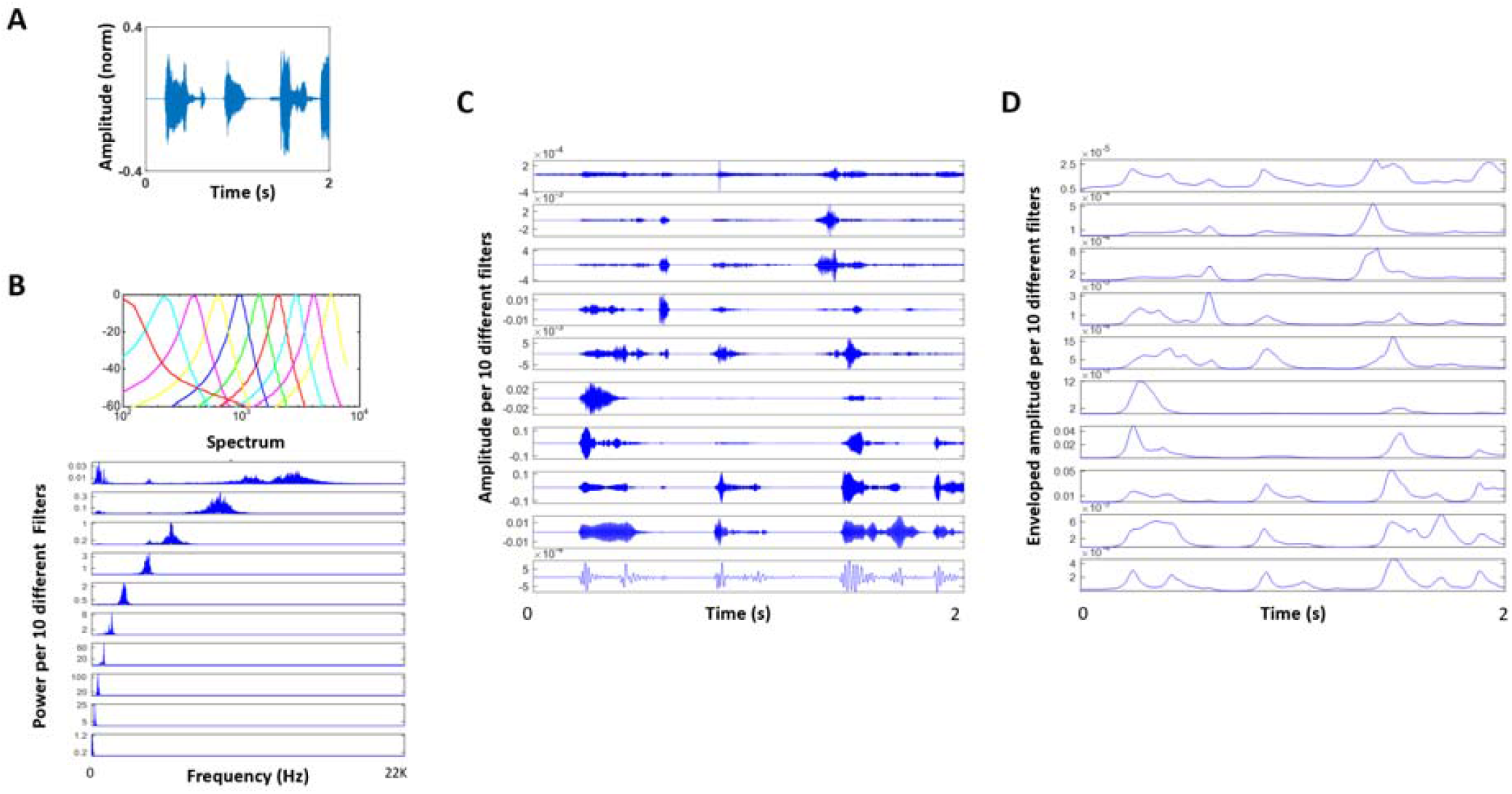
(A) Raw audio waveforms during speech. (B) Gammatone filter to simulate the basilar membrane response. (C) Audio decomposed by Gammatone filter bank. (D) Enveloped audio from (C).

**Figure A2.**
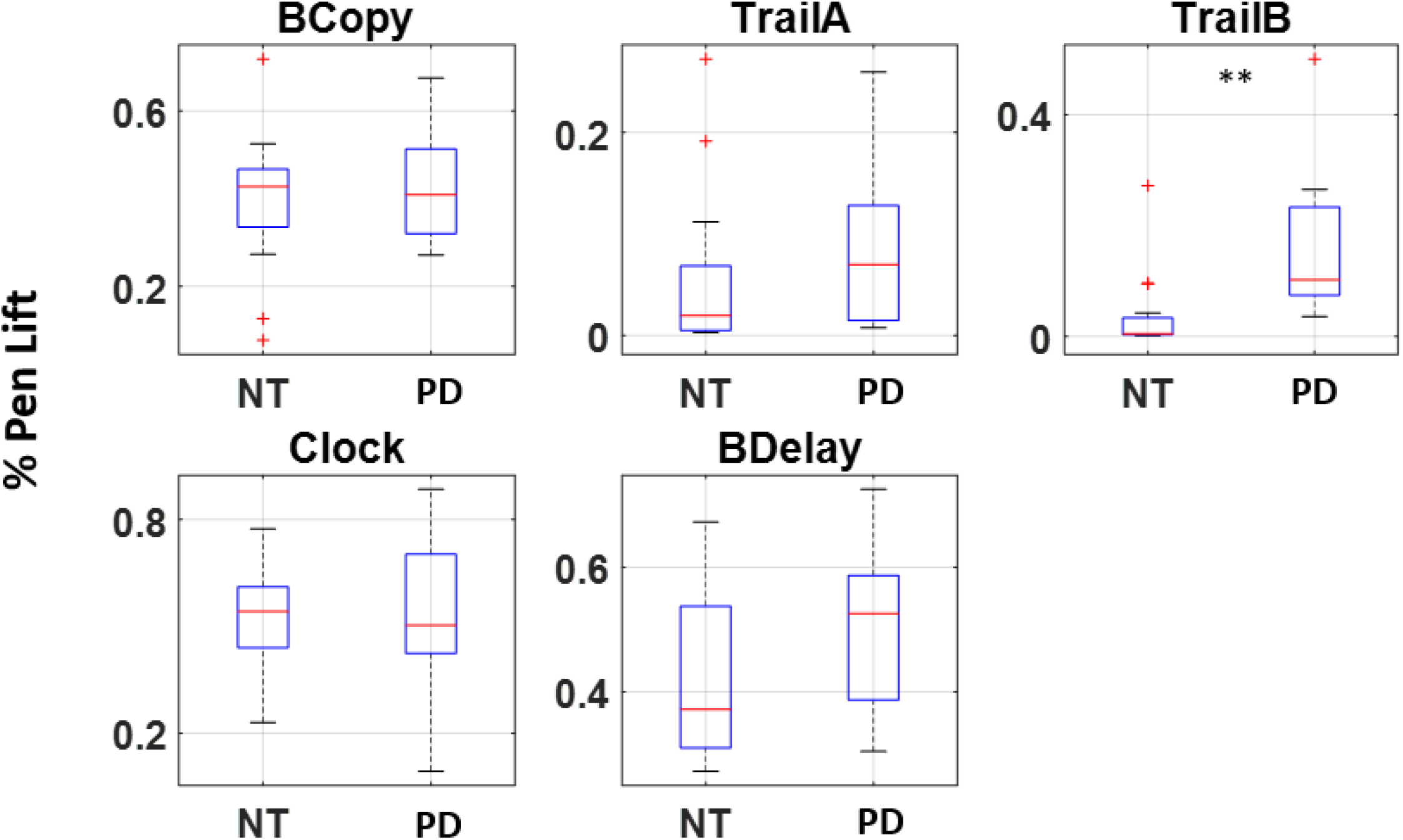
Boxplot of percentage of pen lifts during the entire duration of recording for 5 drawing tasks. ** *p<0.01*

**Figure A3.**
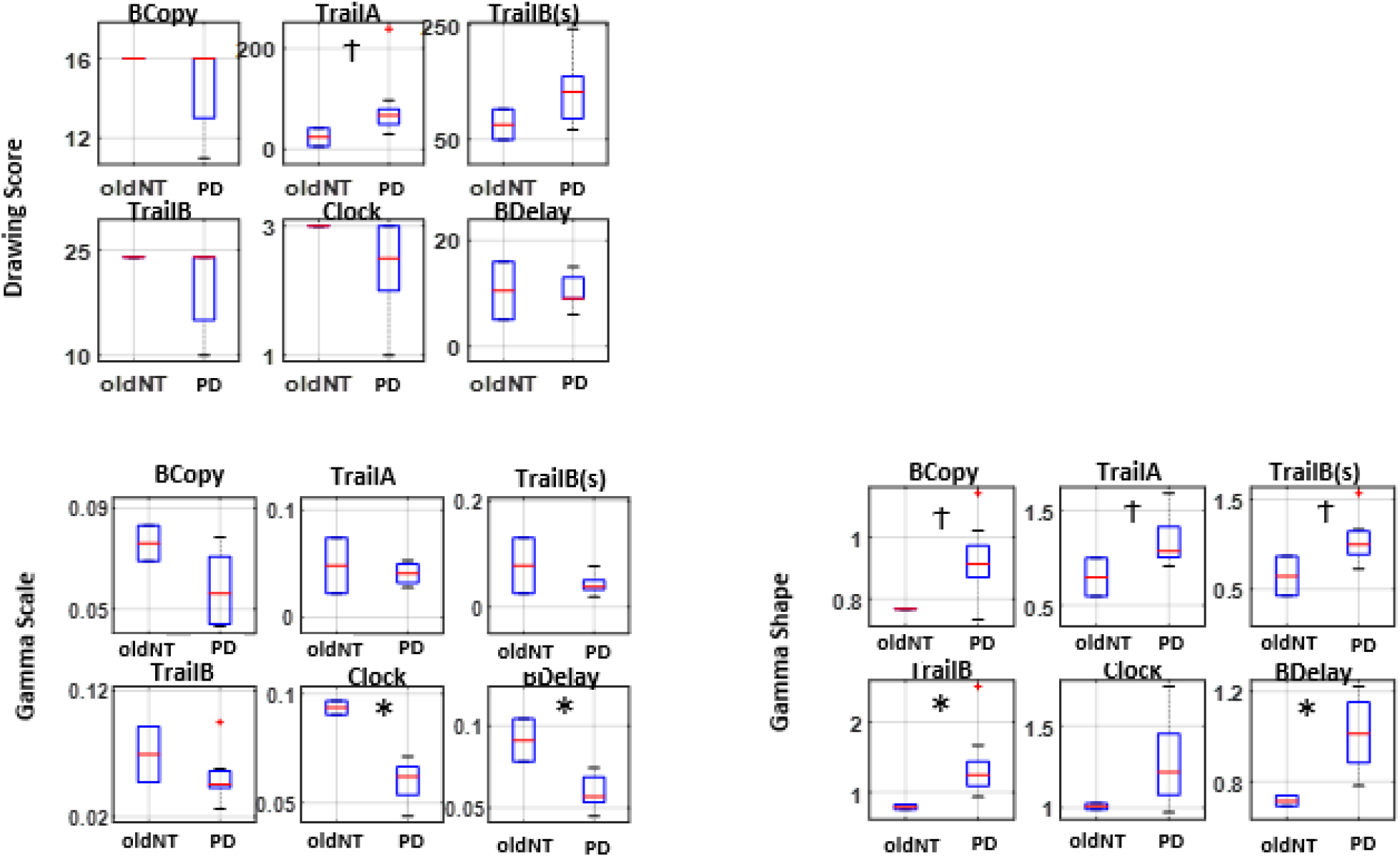
Boxplot of Drawing scores and Gamma parameters between age-matched neurotypical (oldNT) and Parkinson’s patient. * *p<0*.*05*; *t p<0*.*1*

**Figure A4.**
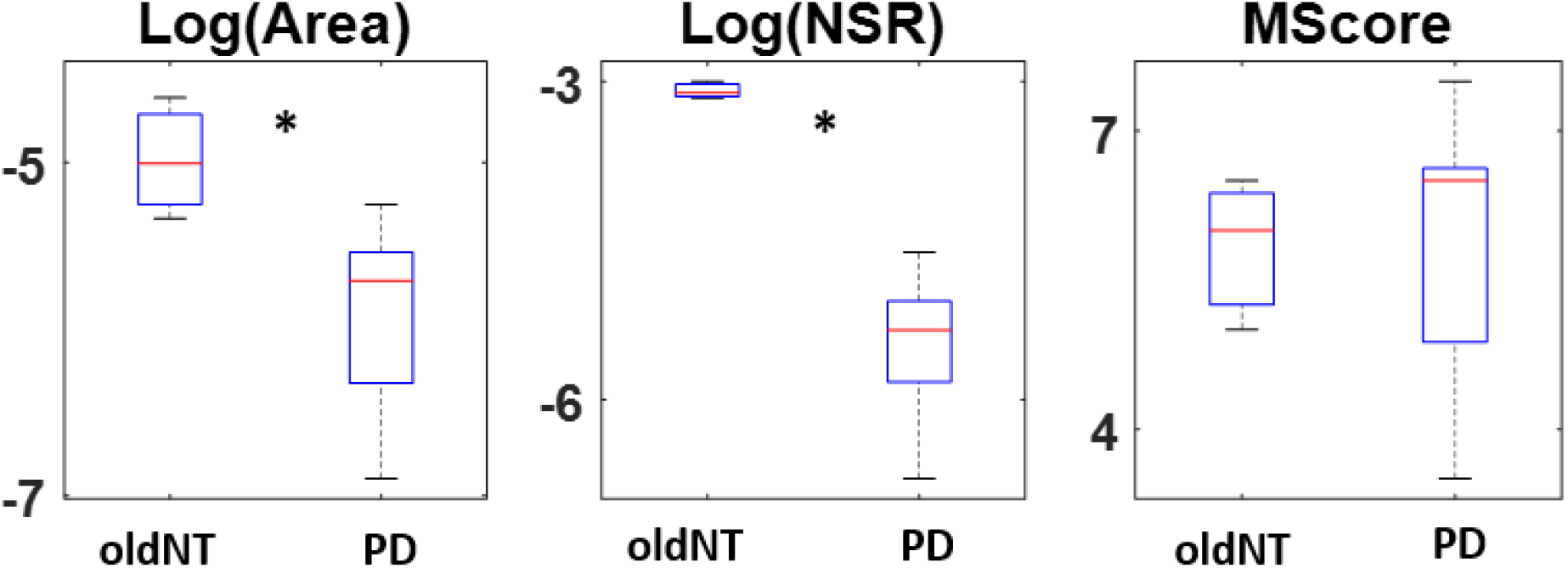
Boxplot logarithm of area, of logarithm of slope NSR, and of memory score between age-matched neurotypical (oldNT) and Parkinson’s patient (PD). * *p<0*.*05*

**Figure A5.**
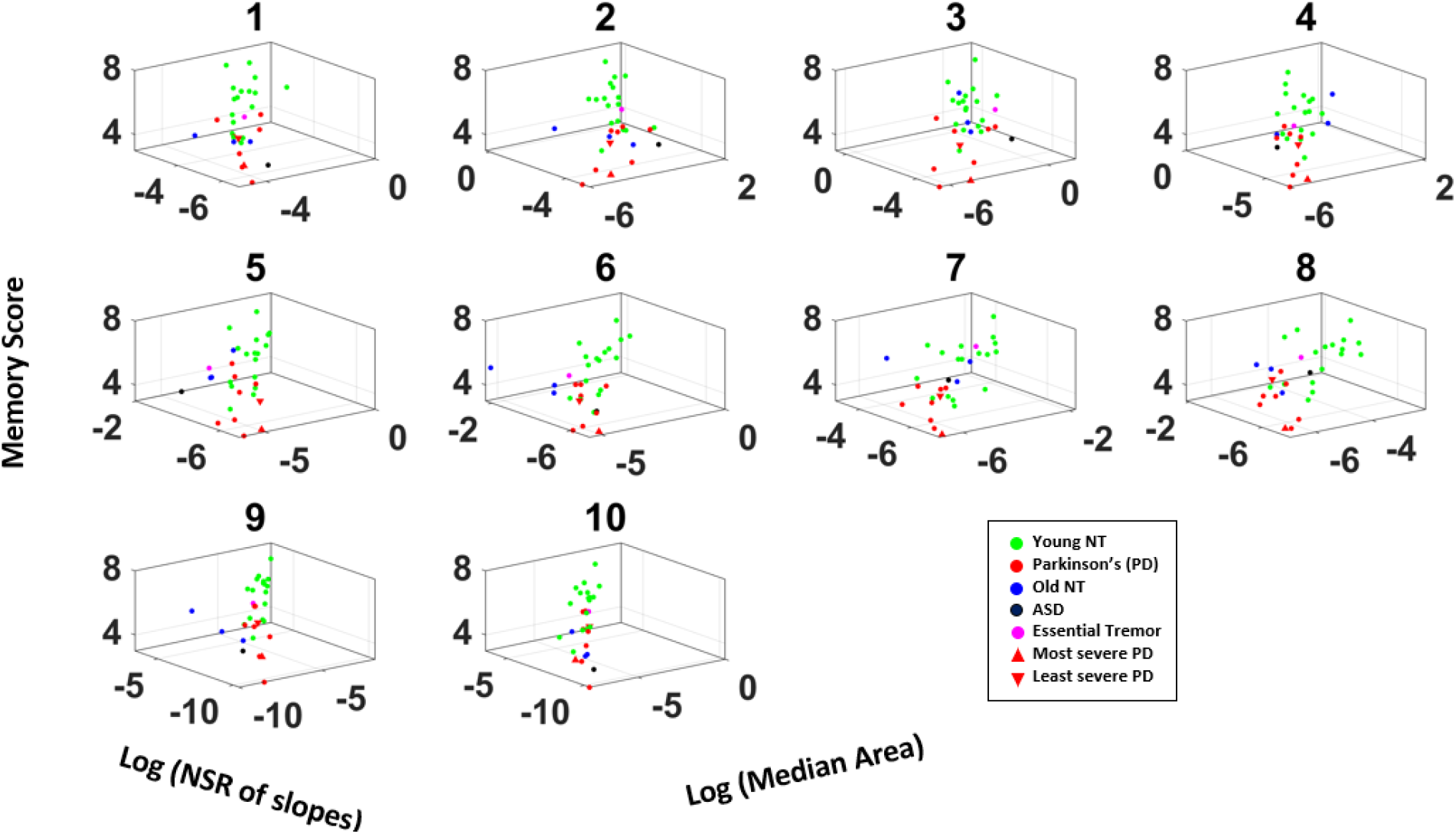
3D plot of memory score, logarithm of slope NSR, and logarithm of median area for all participants, based on differently filtered voice data (#1-10).

**Table A1.** Demographics of patient and age-matched neurotypical participants

| Participant<sup>1</sup> | Gender | Age | UPDRS | Hoehn and Yahr scale | Medication (Motor/Non-Motor)<sup>2</sup> | Pen Data | Voice Data |
| --- | --- | --- | --- | --- | --- | --- | --- |
| PD 1 | M | 51-60 | 6 | 2.5 | Yes/Yes | Yes | Yes |
| PD 2 | F | 61-70 | 44 | 4 | Yes/Yes | Yes | Yes |
| PD 3 | M | 61-70 | 24.5 | 2.5 | Yes/Yes | Yes | Yes |
| PD 4 | M | 61-70 | 19 | 3 | Yes/Yes | Yes | Yes |
| PD 5 | M | 51-60 | 29 | 3 | Yes/Yes | Yes | No |
| PD 6 | F | 61-70 | - | 3 | Yes/No | Yes | Yes |
| PD 7 | M | 61-70 | 16 | 2 | Yes/Yes | Yes | Yes |
| PD 8 | M | 71-80 | 21 | 3 | Yes/No | Yes | Yes |
| PD 9 | F | 71-80 | 27 | 3 | Yes/Yes | Yes | No |
| PD 10 | M | 41-50 | - | 2 | Yes/Yes | Yes | Yes |
| PD 11 | M | 61-70 | 20 | - | - | No | Yes |
| ASD 1 | F | 21-30 | - | - | - | No | Yes |
| ET 1 | M | 31-40 | - | - | - | Yes | Yes |
| Old NT 1 | F | 61-70 | - | - | - | Yes | No |
| Old NT 2 | M | 61-70 | - | - | - | Yes | No |
| Old NT 3 | F | 51-60 | - | - | - | No | Yes |
| Old NT 4 | F | 71-80 | - | - | - | No | Yes |
| Old NT 5 | M | 71-80 | - | - | - | No | Yes |

<sup>1</sup>Participant Abbreviation
PD: Parkinson's disease
ASD: Autism spectrum disorder
ET: Essential tremor
Old NT: age matched neurotypical control
<sup>2</sup> Intake of medication for motor issues / medication for non-motor issues that are known to affect motor performance

**Table A2.** Kruskal-Wallis Test on Parameter Comparison during drawing task

| Parameter | Drawing | yNT<sup>1</sup> vs PD |  | oldNT<sup>2</sup> vs PD |  |
| --- | --- | --- | --- | --- | --- |
|  |  | Chi (1,25) | p-value | Chi (1,10) | p-value |
| Drawing Score | BCopy | 5.80 | 0.02* | 0.72 | 0.40 |
|  | Trail A | 14.95 | 0.00** | 2.95 | 0.09 |
|  | Trail B(s) | 16.54 | 0.00** | 2.26 | 0.13 |
|  | Trail B | 7.63 | 0.01** | 1.05 | 0.31 |
|  | Clock | 6.95 | 0.01** | 1.50 | 0.22 |
|  | BDelay | 7.77 | 0.01** | 0.00 | 1.00 |
| Gamma shape | BCopy | 7.08 | 0.01** | 2.95 | 0.09 |
|  | Trail A | 4.66 | 0.03* | 2.95 | 0.09 |
|  | Trail B(s) | 3.45 | 0.06 | 2.95 | 0.09 |
|  | Trail B | 3.64 | 0.06 | 4.62 | 0.03* |
|  | Clock | 7.80 | 0.01** | 2.26 | 0.13 |
|  | BDelay | 4.31 | 0.04* | 4.50 | 0.03* |
| Gamma scale | BCopy | 6.82 | 0.01** | 2.26 | 0.13 |
|  | Trail A | 0.00 | 1.00 | 0.00 | 1.00 |
|  | Trail B(s) | 0.06 | 0.80 | 0.05 | 0.83 |
|  | Trail B | 0.57 | 0.45 | 1.15 | 0.28 |
|  | Clock | 15.21 | 0.00** | 4.62 | 0.03* |
|  | BDelay | 8.63 | 0.00** | 4.50 | 0.03* |

<sup>1</sup> yNT: Young neurotypical
<sup>2</sup> oldNT: age-matched neurotypical

